# Brain Functional-Structural Gradient Coupling Reflects Development, Behavior and Genetic Influences

**DOI:** 10.1101/2025.09.16.25335918

**Authors:** Simiao Gao, Zhiling Gu, Shengxian Ding, Gefei Wang, Zhengwu Zhang, Hongyu Zhao, Yize Zhao

**Author notes:** These authors contributed equally to this work (Simiao Gao, Zhiling Gu, Shengxian Ding).

## Abstract

Gradients are increasingly used to characterize the brain’s macroscale organization, offering low-dimensional representations of structural and functional connectivity. However, how structural-functional gradient coupling evolves during development and relates to behavioral and molecular features remains unclear. Here, we studied structural-functional gradient coupling across multiple metrics and spatial scales using high-resolution structural and functional connectivity from 7,025 children in the Adolescent Brain Cognitive Development study and 913 adults from the Human Connectome Project. We found that gradient coupling exhibits clear developmental refinement from childhood to adulthood and shows distinct sex-specific patterns. Gradient coupling metrics were significantly associated with a broad range of cognitive and mental health measures and enabled robust out-of-sample prediction under learning methods. Heritability analyses revealed that gradient coupling is strongly influenced by genetic factors. Transcriptomic analyses further demonstrated that highly heritable coupling patterns are enriched for genes expressed in deep-layer excitatory neurons, suggesting that gradient coupling reflects underlying cell-type-specific transcriptional architecture. Together, our findings establish structural-functional gradient coupling as a biologically meaningful feature of brain organization that bridges macroscale connectivity, cognition, behavior, and molecular architecture.

## Introduction

Understanding how the anatomical architecture of the brain supports its functional activity is a central question in neuroscience. Structural and functional connectivity (SC and FC) provide complementary, network-level characterizations of the brain structural and functional organization, where SC captures physical pathways of white matter tracts while FC characterizes temporal synchronization of activity between regions across the whole brain. The relationship between SC and FC, known as structure–function (SF) network coupling, reflects how closely brain functional patterns are shaped by anatomical constraints at a large-scale network level [1]. Early studies investigated global SF network coupling by assuming uniform correspondence across the brain [2, 3]. More recent research has shown that SF network correspondence varied across the cortex, and was higher in sensory-motor regions and lower in the default mode network [3, 4, 5].

Concurrently, recent advances in gradient-based approaches have transformed our understanding of large-scale brain organizations [6]. Rather than characterizing the brain as a set of discrete networks, gradient-based approaches project connectivity patterns into low-dimensional manifolds that reveal continuous axes of organization [7, 8, 9]. Functional gradients consistently map from sensory-motor to association cortex, aligning with microscale cytoarchitecture [10], evolutionary expansion patterns [11], and cognitive hierarchies [6]. Structural gradients capture spatial hierarchies in anatomical architectures, and were shown to progress from locally connected sensorimotor areas to distributed transmodal regions [12]. The spatial alignment between structural and functional gradients offers a more nuanced view of SF coupling, referred to as SF gradient coupling, which captures correspondence between anatomical and functional hierarchies spanning along continuous topography. Despite its promise, SF gradient coupling remains largely understudied. Most research has focused on gradients within a single modality, often limited to functional gradients [6], while the cross-modal correspondence between structural and functional gradients has received limited attention. Even fewer studies have examined how gradient coupling develops over time or varies across individuals. Moreover, it is unclear how gradient coupling varies across individuals and developmental stages, or in relation to behavioral traits or genetic factors. Filling these gaps could offer critical insight into how the brain integrates structure and function over neurodevelopment, how these interactions support cognition and mental health, and how they are shaped by molecular architectures.

Although many aspects of brain development unfold gradually across childhood and adolescence, most studies of SF coupling have focused on either mature adult brains or aggregated developmental trajectories. This leaves a critical gap in understanding how SF alignment differs across key developmental stages, particularly between childhood when neural systems are still forming, and adulthood when brain organizations are more stable and specialized. Previous work has suggested age-related increases in SF coupling during youth, particularly in highly expanded association cortex [13], but no study has yet tested how SF gradient coupling evolves from late childhood through early adulthood, or whether this process differs by sex.

Equally underexplored is the behavioral relevance of SF gradient coupling. While prior research has suggested that SF coupling may support cognitive performance and clinical outcomes [1, 14], the relationship between SF gradient coupling and individual differences in cognition or mental health has rarely been explored. In particular, the potential of SF gradient coupling as a predictive feature for behavioral traits has not been investigated. It remains unclear whether variation in the alignment of structural and functional hierarchies can explain altered cognition and behavior, and if so, which coupling patterns are most informative. Addressing this gap is critical for advancing our understanding of how macroscale brain organization supports complex traits and for identifying potential neuromarkers to inform neurodevelopmental and psychiatric conditions.

Furthermore, both SC and FC are known to be heritable [15, 16, 17], and emerging evidence also suggests that SF coupling was also shaped by genetic factors [14, 18]. However, it remains unclear whether the alignment between structural and functional gradients is heritable, which brain systems are most susceptible to genetic influence, or how such effects vary across cortical hierarchies. Meanwhile, transcriptomic atlases such as the Allen Human Brain Atlas (AHBA) now enable researchers to examine whether neuroimaging-derived phenotypes reflect underlying cell-type–specific gene expression patterns [19]. If spatial variations in gradient coupling correspond to distinct molecular signatures, it would help provide crucial biological context for interpreting this emerging marker for brain organization.

To address these gaps, in this work, leveraging multi-dimensional data from 7,025 children in the Adolescent Brain Cognitive Development (ABCD) study and 913 adults from the Human Connectome Project (HCP), we constructed brain SF gradient coupling under various metrics and spatial scales across development stages, and comprehensively uncovered its correspondence with cognitive and behavioral domains, as well as genetics and molecular architectures. Our approaches and results revealed that SF gradient coupling undergoes systematic neurobiological refinement from late childhood to early adulthood with distinct sex-specific trajectories. We further demonstrated that gradient coupling robustly predicts a wide range of cognitive and mental health traits across cohorts and methods, establishing it as a behaviorally meaningful neuromarker. Extending beyond phenotype, we showed that brain gradient coupling is highly heritable, especially in transmodal and sensorimotor networks. Finally, we integrated transcriptomic data from the AHBA to reveal that heritable gradient coupling patterns are enriched for genes expressed in deep-layer excitatory neurons, linking macroscale brain organization to cell-type-specific molecular architecture. Together, these findings positioned SF gradient coupling as a biologically grounded, developmentally refined, and behaviorally relevant feature of human brain organization.

## Results

### Gradient coupling reveals developmental refinement across cortical hierarchies

Leveraging multi-modal raw neuroimaging data spanning childhood (*n* = 7, 938) to adulthood (*n* = 944), we constructed fine-scale structural and functional connectivity for each subject from T1-weighted, diffusion, and functional magnetic resonance imaging (MRI) data. We then derived individual-level cortical structural and functional gradients, and constructed both macroscale-level and subnetwork-level SF gradient couplings as illustrated in Fig. 1. The detailed data processing and metrics construction procedures are described in the Methods section.

**Fig. 1.**
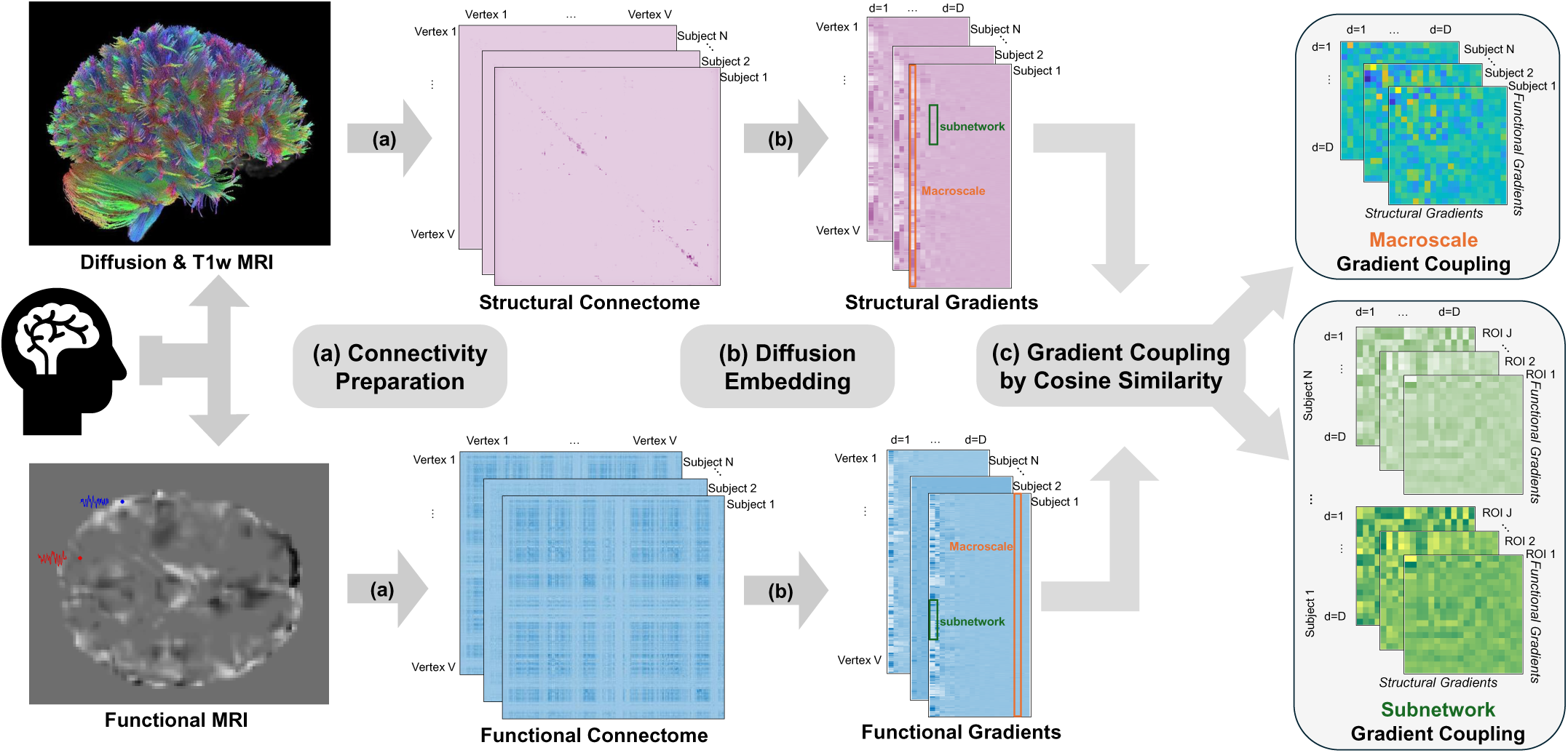
Overview of SF gradient coupling construction. **a** Preprocessed diffusion MRI, T1-weighted MRI, and resting-state functional MRI were processed using the Surface-Based Connectivity Integration pipeline to generate individual-level structural and functional connectomes. **b** Functional and structural gradients were then extracted from the resulting connectivity matrices using diffusion embedding via the BrainSpace toolbox, yielding low-dimensional representations of cortical organization. **c** Gradient coupling was quantified at both the macroscale and subnetwork levels using cosine similarity between corresponding functional and structural gradients.

To investigate SF gradient coupling and its developmental dynamics, we first examined the architectures of structural and functional gradients separately and assessed their age-related changes. Although certain cortical regions showed high connectivity strength in both structural and functional domains (Supplementary Fig. 1), the corresponding gradients revealed distinct organizational principles: structural gradients primarily reflected the topology of long-range white matter pathways, whereas functional gradients delineated the segregation of functional subnetworks. Visualization of these gradients across cohorts was provided in Fig. 2a, b, g, h, illustrating how their spatial layout differed and evolved with age. In children, the principal functional gradient predominantly captured a sensorimotor-to-visual axis, reflecting the early organization of primary systems. Their second functional gradient began to resemble the adult principal axis, indicating the gradual emergence of transmodal association networks. In contrast, adults exhibited a well-defined principal functional gradient extending from primary sensorimotor areas to transmodal hubs, consistent with the mature hierarchical organization of the cortex. Structural gradients showed even more pronounced developmental differences. In children, the primary structural gradient separated visual and somatomotor systems but displayed strong asymmetry along the second gradient, consistent with ongoing white matter maturation and lateralized development supporting functions such as language and attention. In adults, the primary gradient also separated visual and somatomotor systems but exhibited greater bilateral symmetry and spatial precision, indicating refined sensory network segregation. Collectively, they highlighted the evolving divergence and convergence of structural and functional gradients during development, motivating our investigation of SF gradient coupling as a biologically interpretable measure of alignment between anatomical and functional brain organization across development.

**Fig. 2.**
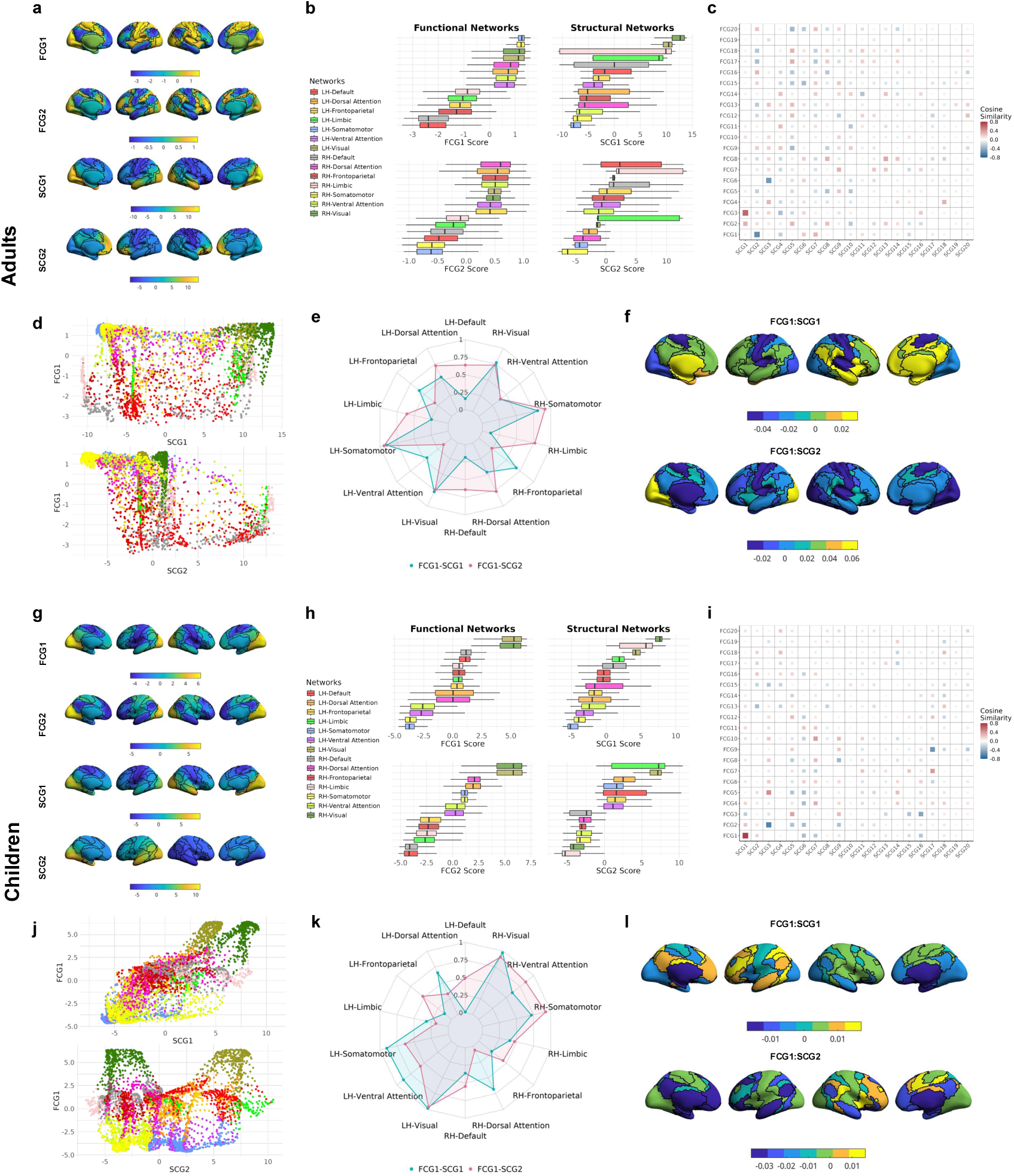
Structural and functional gradients and their couplings at macroscale and subnetwork levels in adults (a–f) and children (g–l). a, **g** Principal and secondary gradients derived from cohort-level functional connectivity (FCG1, FCG2) and structural connectivity (SCG1, SCG2), displayed on the cortical surface with color scales indicating gradient values. **b, h** Distributions of gradient values across the Yeo-7 subnetworks, presented as boxplots and ordered by median gradient value. **c, i** Pairwise cosine similarity matrices between the top 20 structural and functional gradients. Structural gradients (SCG1∼SCG20) are shown along the x-axis and functional gradients (FCG1∼FCG20) along the y-axis. **d, j** Scatter plots of FCG1 versus SCG1 and SCG2, illustrating examples of strong and weak macroscale gradient coupling. Color coding matches the networks shown in panels **b** and **h**. **e, k** Radar plots showing the absolute values of gradient coupling (FCG1:SCG1 and FCG1:SCG2) across Yeo-7 networks. **f, l** Differences in Yeo-7 subnetwork-level gradient coupling (FCG1:SCG1 and FCG1:SCG2) between males and females, shown as the mean coupling in males minus that in females for each subnetwork.

The spatial correspondence between functional and structural gradients reflected a core principle of structure–function association across cortical hierarchies. To quantify this correspondence, we computed pairwise cosine similarity between the cohort-level structural and functional gradients, resulting in macroscale SF gradient coupling matrices (Fig. 2c, i). Throughout this article, we denote SF gradient coupling between the the *i*th functional gradient (FCG) and the *j*th structural gradient (SCG) as FCG*i*:SCG*j*. In children, a strong coupling was observed between the principal functional and structural gradients (SF gradient coupling *c* = 0.77 for FCG1:SCG1, *p <* 10^−6^), suggesting that early functional organization, particularly along the sensorimotor-to-visual axis, remains closely aligned with the underlying structural scaffold. In contrast, the same gradient pair in adults showed much weaker coupling (*c* = 0.04, *p* = 0.0064). We further observed that the SF gradient coupling strength between the principal functional gradient and the second structural gradient (FCG1:SCG2) increased from near-zero levels in children to a robust level in adults.

Having established the macroscale gradient coupling strength across different gradient pairs and development stages, we next examined how SF gradient coupling patterns manifested within individual functional systems, as defined by the Yeo-7 canonical subnetworks [20]. We observed substantial heterogeneity in SF gradient alignment at the subnetwork level: some networks exhibited strong coupling even when global alignment was weak, whereas others displayed decoupling despite strong global coupling. To illustrate this coupling heterogeneity, we focused on two representative gradient pairs, FCG1:SCG1 and FCG1:SCG2, each showing strong alignment in one age group but weak alignment in the other (Fig. 2c, d, i, j). Notably, strong coupling was consistently observed in primary sensory and motor networks, whereas association networks such as frontoparietal and default mode systems showed greater decoupling. This pattern was consistent with prior findings of reduced structure and function correspondence in transmodal regions [21]. To further assess these network-specific patterns, we visualized coupling strength across networks in each hemisphere using radar plots (Fig. 2e, k). The results highlighted relatively high and consistent SF gradient alignment in unimodal systems across developmental stages, in contrast to the weaker and more variable coupling observed in higher-order networks. Together, these findings established macroscale and subnetwork-level SF gradient coupling as two equally informative metrics for quantifying correspondence between brain structural and functional organization. We therefore included both metrics in downstream analyses to assess their associations with behavioral, demographic, and genetic factors.

### Gradient coupling and mental health associations by age and sex

We assessed the associations between SF gradient coupling and a broad range of behavioral and demographic outcomes at both the macroscale and subnetwork levels. To ensure robust and generalizable estimates, we implemented two predictive modeling approaches: kernel ridge regression (KRR) [22] and multilayer perceptrons (MLP) [23], with 100 random train-test splits. Detailed procedures for model fitting and evaluation are provided in the Methods section.

Aligned with previous findings that SF coupling is associated with psychiatric vulnerability [24], our results revealed modest associations between gradient coupling and mental health outcomes, with slightly stronger effects observed in adults. The strongest association (Pearson’s *r* = 0.11) was observed for externalizing symptoms (External, Fig. 3a). Furthermore, the association between SF gradient coupling and outcomes varied by scale and age cohort: in adults, subnetwork-level SF gradient coupling showed stronger association with psychiatric symptoms than macroscale gradient coupling, whereas in children, macroscale SF gradient coupling yielded exhibited associations. This divergence suggested that the spatial scale at which gradient coupling was behaviorally relevant may shift across development.

**Fig. 3.**
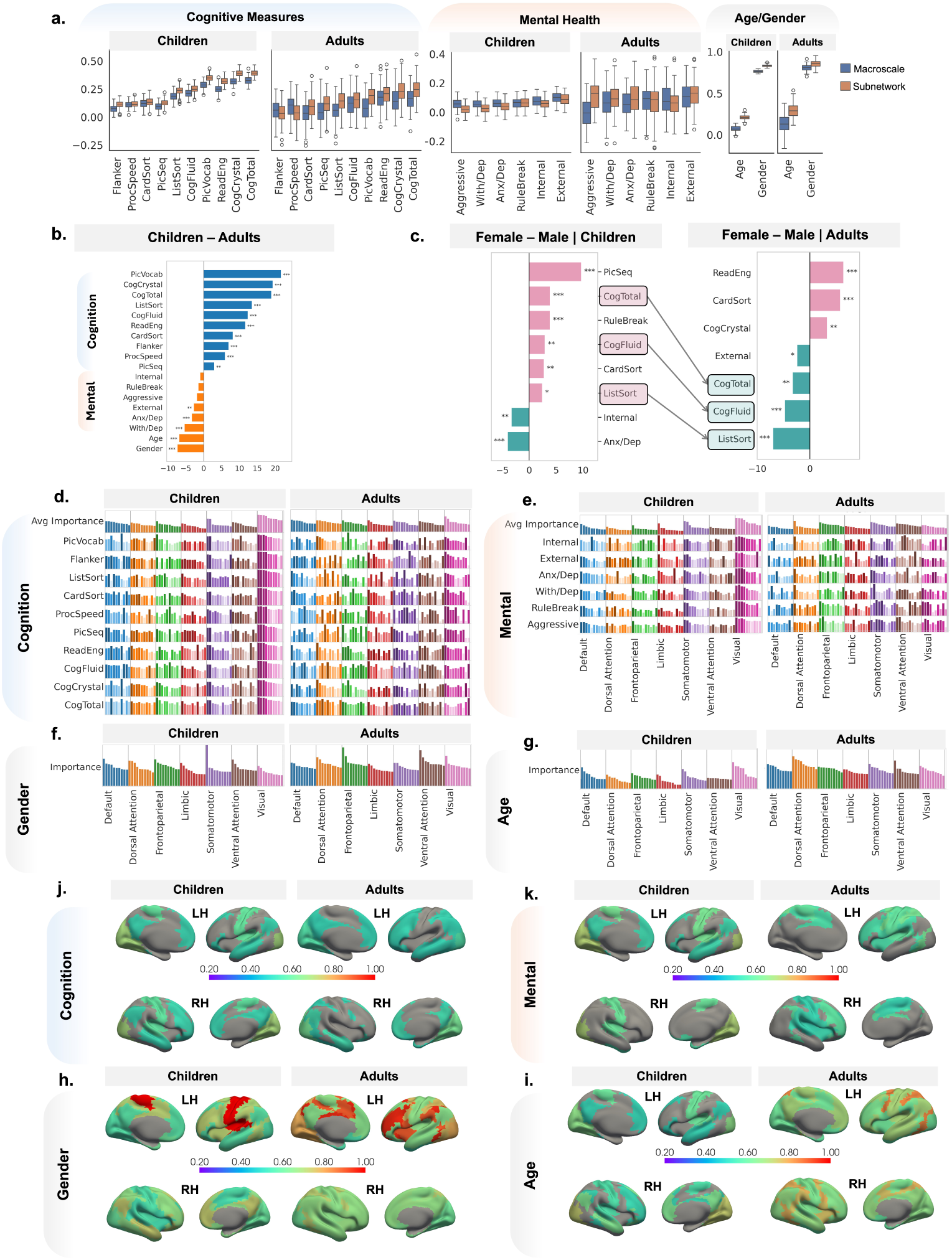
The association between SF gradient couplings and behavioral outcomes and demographics. **a** Boxplots illustrating the association between gradient coupling and behavioral outcomes at the macroscale and Yeo-7 subnetwork levels. Correlation coefficients are shown for continuous measures (cognitive, mental health, and age), and AUC values are displayed for gender. **b, c** Differences in gradient coupling-behavior associations across cohorts and genders. The x-axis represents t-statistics; asterisks denote significance levels (^∗^ : *p <* 0.05, ^∗∗^ : *p <* 0.01, ^∗∗∗^ : *p <* 0.001). **d-g** Feature importance of top subnetwork-level gradient coupling metrics for predicting individual behavioral outcomes (lower panels) and domain-averaged importance (upper panels). Darker and taller bars indicate greater predictive importance. **j-i** The highest averaged feature importance for gradient coupling within each subnetwork in predicting domain outcomes.

To formally assess cohort-related differences, we conducted two-sample *t* -tests comparing coupling–outcome association strengths between children and adults. As shown in Fig. 3b, gradient coupling was more strongly associated with mental health outcomes in adults. While batch or dataset effects cannot be ruled out, this pattern may reflect developmental shifts in the brain’s vulnerability architecture, wherein deviations in gradient coupling gain behavioral significance during a period of increased susceptibility to psychiatric disorders [25].

Beyond mental health, gradient coupling also showed predictive utility for sex classification, with area under the curve (AUC) values of 0.80 in children and 0.83 in adults (Fig. 3a). This aligned with previous findings of sex-related differences in SF coupling patterns [26]. Visual inspection of gradient coupling topographies revealed distinct spatial patterns between males and females (Fig. 2f, l). For instance, in the FCG1:SCG1 pair, sex differences in gradient coupling were more pronounced within association networks in children, whereas in adults, they were more evident in unimodal regions. These spatial and developmental shifts suggested that SF gradient coupling captured a fundamental axis of sex-dimorphic brain architecture. Our findings were consistent with prior theories of sexually differentiated maturation trajectories [27] and supported the utility of gradient coupling as a biologically grounded marker for modeling sex-specific brain–behavior relationships.

To explore how these sex and cohort effects interact in shaping the mental health relevance of gradient coupling, we further stratified the cohorts by sex and re-estimated coupling–outcome associations (Fig. 3c). In children, rule-breaking behavior (RuleBreak) showed a significant sex difference, with females exhibiting a stronger association with gradient coupling. Conversely, males showed greater associations for internalizing (Internal) and anxiety/depression (Anx/Dep) symptoms. In adults, however, the only significant sex difference was for externalizing symptoms, where stronger associations were observed in males. These findings suggested that the psychiatric correlates of gradient coupling were modulated by both developmental stage and sex.

### Cognitive relevance of gradient coupling varies across development and sex

Previous studies reported significantly altered SC and FC between hippocampal subregions, implicated in cognition and emotion, and cortical areas in individuals with cognitive impairment [28, 29]. Extending these findings, [30] demonstrated that while SF coupling was largely preserved at the whole-brain level in mild cognitive impairment, it was selectively increased in the dorsal attention network and decreased in the ventral attention network. These findings suggested that SF gradient coupling may reflect compensatory or pathological reorganization and could serve as a predictor of cognitive outcomes.

Building on this, we evaluated the relationship between SF gradient coupling and cognitive function across development. Both macroscale and subnetwork-level gradient coupling showed broadly similar association patterns, with subnetwork-level coupling showing slightly stronger associations with most cognitive outcomes (Fig. 3a). Remarkably, the strength of the association between gradient coupling and cognitive performance was greatest in children (*r* = 0.22, *p <* 10^−6^), particularly for integrative cognitive scores such as the Total Cognition Composite (CogTotal, *r* = 0.36, *p <* 10^−6^), and became weaker in adults (*r* = 0.20, *p <* 10^−6^). The *t*-statistics in Fig. 3b further confirmed that gradient coupling was more predictive of cognitive outcomes in children, underscoring the greater behavioral relevance of SF gradient alignment during this critical developmental stage. This developmental pattern aligned with prior work suggesting that adolescence was a period of heightened neural plasticity and large-scale network reorganization [31, 32], during which structural and functional hierarchies may become increasingly aligned to support emerging executive abilities [13].

Having observed sex differences in the associations between gradient coupling and mental health, we next examined whether similar sex-dependent patterns exist for cognitive outcomes across development. As shown in Fig. 3c, in children, females exhibited stronger coupling–cognition associations for CogTotal, Fluid Cognition Composite (CogFluid), as well as individual domains including Picture Sequence Memory (PicSeq), Dimensional Change Card Sort (CardSort), and List Sorting Working Memory (ListSort). In contrast, in adults, males showed stronger associations with measures such as CogFluid, CogTotal, and ListSort, while CardSort remained more strongly linked to gradient coupling in females. This developmental reversal may reflect sex-specific trajectories of brain maturation, with males having been shown to reach peak brain volumes later than females [33]. Together, these findings pointed to a dynamic interaction between SF gradient coupling and cognitive functions, shaped by both developmental stage and sex.

### Key gradient couplings exhibit a shift from unimodal to transmodal regions across developmental stages

To identify the neural circuits underlying associations between SF gradient coupling and behavioral and demographic outcomes, we analyzed the feature importance of subnetwork-level gradient coupling measures. For both cognitive (Fig. 3d, j) and mental health outcomes (Fig. 3e, k), we observed a clear developmental shift in the spatial distribution of predictive subnetwork features. In children, unimodal regions, particularly the somatomotor and visual networks, contributed most strongly to the prediction of both cognitive and mental health outcomes. In contrast, adults showed increased involvement of transmodal regions, including the frontoparietal and ventral attention networks, suggesting a maturation-related transition from sensory-driven to integrative functional systems.

A similar cohort-related redistribution emerged in associations with sex (Fig. 3f, h) and age (Fig. 3g, i). In children, predictive gradient coupling features for sex were primarily localized in the somatomotor network, whereas in adults, these features shifted toward the frontoparietal, ventral attention, and visual systems, indicating a dynamic reorganization of subnetwork-level coupling patterns across development. For coupling-age association, children relied more heavily on the default mode, somatomotor, and visual networks, while in adults, predictive contributions came increasingly from the dorsal and ventral attention networks.

These findings aligned with prior evidence that gradient-based organization was more localized in unimodal cortices during early developmental stages [34]. This evolving functional architecture was further reflected in Fig. 3a, where coupling–behavior associations exhibited lower inter-individual variability in children relative to adults across all behavioral domains. Taken as a whole, these results suggested that SF gradient coupling became increasingly differentiated and individualized with age, offering a potential neural substrate for behavioral diversity in adulthood. Additionally, as shown in Supplementary Figs. 3 and 4, we observed consistent overall patterns in the feature importance maps, supporting the robustness of our analysis and the reliability of the proposed feature importance metrics.

### Gradient coupling exhibits distinct and hierarchically organized genetic contributions

We further investigated how genetic factors shape SF gradient coupling across developmental stages by estimating the heritability of each coupling metric. In addition to examining heritability within Yeo-7 canonical subnetworks, we also computed heritability for gradient coupling defined at a finer spatial resolution of the Desikan–Killiany (D-K) atlas [35], which comprised 34 cortical regions per hemisphere. Including the D-K atlas allowed us to more precisely characterize the spatial distribution of heritability across the brain, and to directly compare these patterns with the spatial organization of the gradient coupling itself. As shown in Fig. 4a, c, j, l, the average gradient coupling strength matrices for both children and adults displayed a weak diagonal trend, with stronger alignment among lower-order gradients. However, this pattern was diffuse and lacked a consistent decay across gradient orders. In contrast to the non-uniform decay in coupling strength, the heritability of these gradient coupling metrics revealed a more explicit and systematic hierarchy, where heritability estimates decreased monotonically with increasing gradient coupling in both cohorts and parcellation schemes (Fig. 4b, d, k, m). This pattern was particularly pronounced under the D-K atlas, likely due to its finer spatial resolution that enhanced the capture of localized, region-specific genetic signals. Importantly, this hierarchical decline in heritability was not simply a byproduct of weaker coupling strength at higher gradient orders. Rather, it reflected an underlying gradient-based genetic hierarchy, suggesting that lower-order SF gradient couplings, presumed to capture core axes of SF brain organization, were more strongly constrained by genetic factors.

**Fig. 4.**
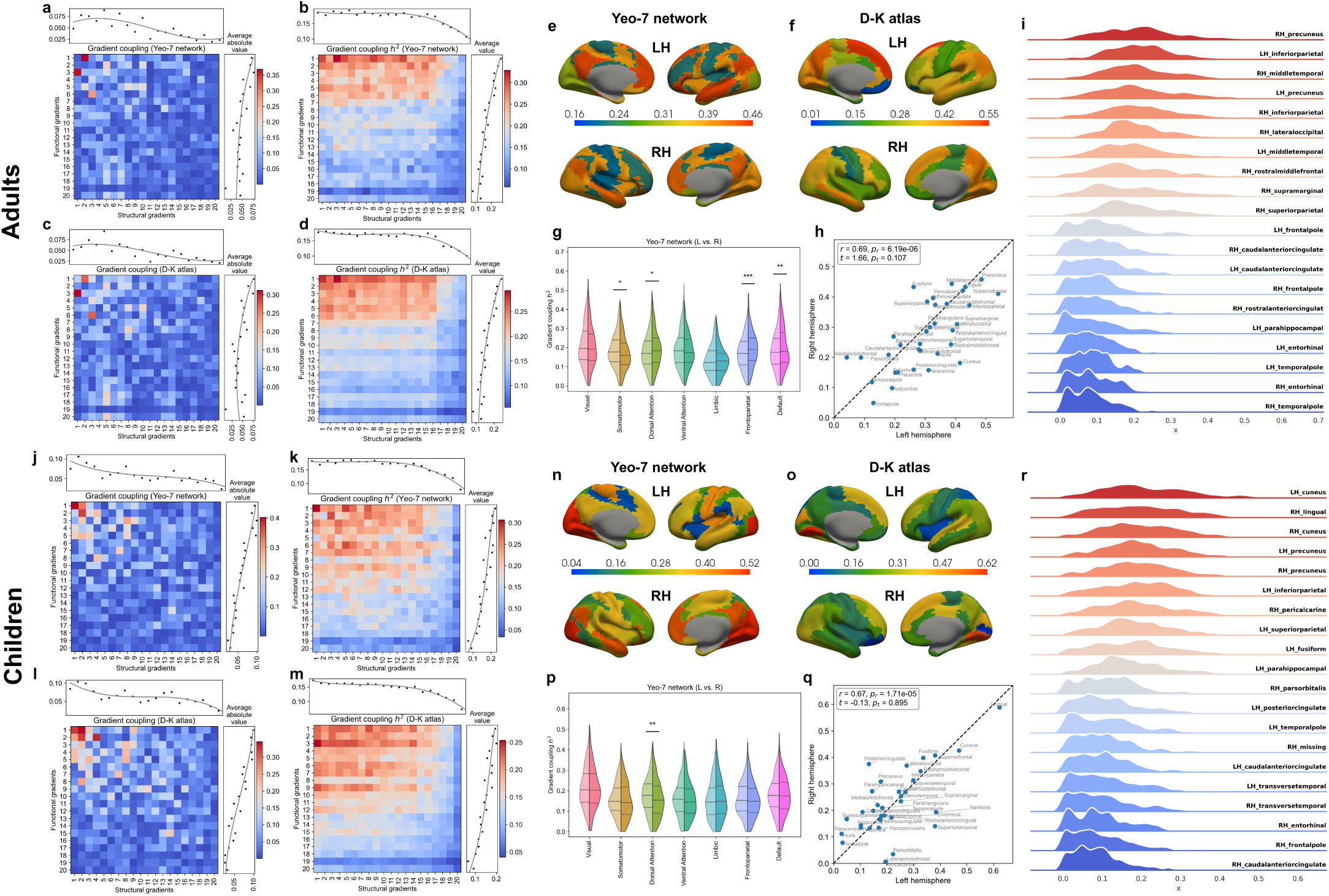
Heritability analyses of gradient coupling. **>a-d, j-m** Subnetwork-level gradient coupling and its heritability in the two cohorts, averaged across the Yeo-7 subnetworks (**a, b, j, k**) and D-K atlas (**c, d, l, m**) respectively. **e, n** Heritability estimates of Yeo-7 subnetwork-level coupling between FCG1 and SCG1. **f, o** Heritability estimates of D-K atlas-level coupling between FCG1 and SCG1. **g, p** Distribution of heritability for Yeo-7 subnetwork-level gradient coupling, separated by hemispheres. Asterisks denote significance based on Bonferroni-corrected pairwise t-tests (^∗^ : *p <* 0.05, ^∗∗^ : *p <* 0.01, ^∗∗∗^ : *p <* 0.001). **h, q** Heritability of the D-K atlas-level gradient coupling between FCG1 and SCG1, grouped by hemisphere, along with corresponding Pearson correlations (*r*), t-statistics, and p-values (*p_r_* and *p_t_* respectively). **i, r** Distribution of heritability estimates for D-K atlas-level gradient coupling.

We also observed that the heritability of SF gradient coupling was broadly consistent across developmental stages, with slightly higher averages in adults (Yeo-7: *h*^2^ = 17.00% versus 16.27%; D-K: *h*^2^ = 16.00% versus 14.49%). The highest heritability estimates for individual subnetwork-level gradient coupling reached 62.75% in adults and 62.19% in children. At the macroscale gradient coupling, the mean heritability increased further to 22.12%, with maximum values up to 60.82%. To contextualize these estimates, we compared them to the heritability of unimodal functional and structural gradients (Supplementary Fig. 5). Functional gradients showed moderate heritability in both cohorts. In adults, the heritability for the first three gradients averaged *h*^2^ = 23.49%, 20.86%, and 17.83%, with corresponding maxima of 66.86%, 54.31%, and 60.61%. Comparable estimates were observed in children (mean: 23.97%, 23.28%, and 17.51%; max: 51.95%, 57.24%, and 42.39%). In contrast, structural gradients were consistently more heritable, especially in adults, reaching mean heritability for the first three gradients 48.12%, 33.77%, and 39.72% (max: 65.59%, 60.48%, and 65.88%). However, in children, structural gradient heritabilities were notably lower (mean: 32.16%, 30.77%, and 22.91%; max: 48.02%, 67.58%, and 57.59%). While the mean heritability of SF gradient coupling was comparable to that of unimodal gradients, its maximum values, particularly for those defined under D-K atlas in children, exceeded those of any individual functional or structural gradient. These findings suggested that SF gradient coupling captured highly heritable multimodal axes of brain organization that were not fully explained by either modality alone. Moreover, stronger heritability in children suggested that genetic influences on SF gradient coupling played a key role in early brain network development.

### Gradient coupling reveals regionally distinct and developmentally specific genetic topographies

Building on the hierarchical heritability patterns across gradient indices, we further examined the spatial topography of SF gradient coupling heritability across the cortex. We focused on the coupling between the first structural and functional gradients (FCG1:SCG1), which consistently exhibited high genetic influence in both cohorts. We first analyzed the subnetwork-level organization of coupling heritability using the Yeo-7 functional networks (Fig. 4e, n, g, p). In adults, the default mode network showed the highest heritability in both hemispheres, followed by the dorsal attention network in the left hemisphere and the visual network in the right. In children, heritability peaked in the visual and somatomotor networks in the left hemisphere and in the visual and default mode networks in the right. Across cohorts, the visual and default mode networks consistently showed the highest median heritability, while the limbic network ranked lowest. These findings indicated that genetic influences on SF gradient coupling spanned both unimodal and transmodal systems, with a greater emphasis on sensory networks during childhood and more distributed, higher-order engagement in adulthood. To localize these effects more precisely, we examined the regional coupling heritability using the D-K atlas (Fig. 4f, o, i, r). Regions mapping to the visual and default mode systems again showed the highest heritability across cohorts, consistent with the subnetwork-level results. Specifically, within the default mode network, the posterior and rostral anterior cingulate and precuneus were among the most heritable in both cohorts, with stronger effects in adults, suggesting increasing genetic regulation of transmodal hubs during maturation. In contrast, within the visual network, the lateral occipital cortex, cuneus, and lingual gyrus showed the highest heritability across development, with more pronounced effects in children, reflecting stronger genetic control over early-developing sensory systems. Children also exhibited elevated heritability in somatomotor and ventral attention regions, such as the precentral gyrus, insula, and bankssts, further supporting the developmental specificity of genetic influence on structural-functional alignment.

To assess hemispheric patterns, we conducted correlation analyses of regional heritability estimates between hemispheres (Fig. 4h, q). Heritability estimates between hemispheres were highly correlated and symmetric in both cohorts (adults: *r* = 0.69*, p_r_* = 6.19 × 10^−6^; children: *r* = 0.67*, p_r_* = 1.71 × 10^−5^), with no significant lateralization detected (adults: *p_t_* = 0.11; children: *p_t_* = 0.90). In children, bilateral symmetry was especially strong, with regions like the lingual gyrus and cuneus consistently ranking among the most heritable in both hemispheres, while slightly higher heritability appeared in the cuneus and superior parietal lobule in the left hemisphere. In adults, greater inter-hemispheric divergence was observed: the left superior frontal gyrus (*h*^2^ = 53.97%) and the right precuneus (*h*^2^ = 45.82%) showed the highest regional heritability within their respective hemispheres, and broader patterns showed divergence between frontal (left) and occipital/prefrontal (right) regions. These asymmetries could indicate a developmental transition from symmetric, sensory-dominant heritability in childhood to more lateralized, control-oriented organization in adulthood.

### Transcriptomic signatures highlight the cellular basis of SF gradient coupling heritability

To explore the cellular basis of heritability variation in SF gradient coupling, we conducted imaging transcriptomics analyses by integrating D-K atlas-level heritability estimates with human cortical gene expression data obtained from the AHBA [38] and cell-type annotations from single-cell RNA sequencing (scRNA-seq) data of the human cortex [37]. The scRNA-seq reference included 20 brain cell types (Fig. 5a), enabling cell-type-specific enrichment analyses of genes associated with the heritability of gradient coupling, as well as spatial correlation analyses of genes associated with regional heritability of SF gradient coupling. We focused on the heritability of SF gradient coupling between the first structural and functional gradients (FCG1:SCG1), as this pair reflected the principal axes of cortical organization, exhibited the strongest SF gradient alignment, and showed the strongest genetic influence across cohorts. The D-K atlas was selected for its anatomical resolution and compatibility with AHBA, enabling region-wise integration of imaging genetics and transcriptomic data. Gene set enrichment analyses (GSEA, [39]), performed across 66 cortical regions with 15,633 gene expression profiles ranked by their association with regional coupling heritability, revealed significant enrichment for excitatory neuron subtypes in both adults and children (Fig. 5b, c). Particularly strong signals were observed for excitatory neurons 3 and 4, corresponding to pyramidal neurons located in deep cortical layers (layers 5–6) implicated in cortico-cortical communication [37], which was consistent with the underlying neurobiological substrates of SF gradient coupling. Notably, this enrichment profile remained highly stable when repeating the analysis using the average heritability across all SF gradient coupling metrics, indicating robust and widespread contributions of excitatory neurons to genetic regulation of multimodal connectivity alignment.

**Fig. 5.**
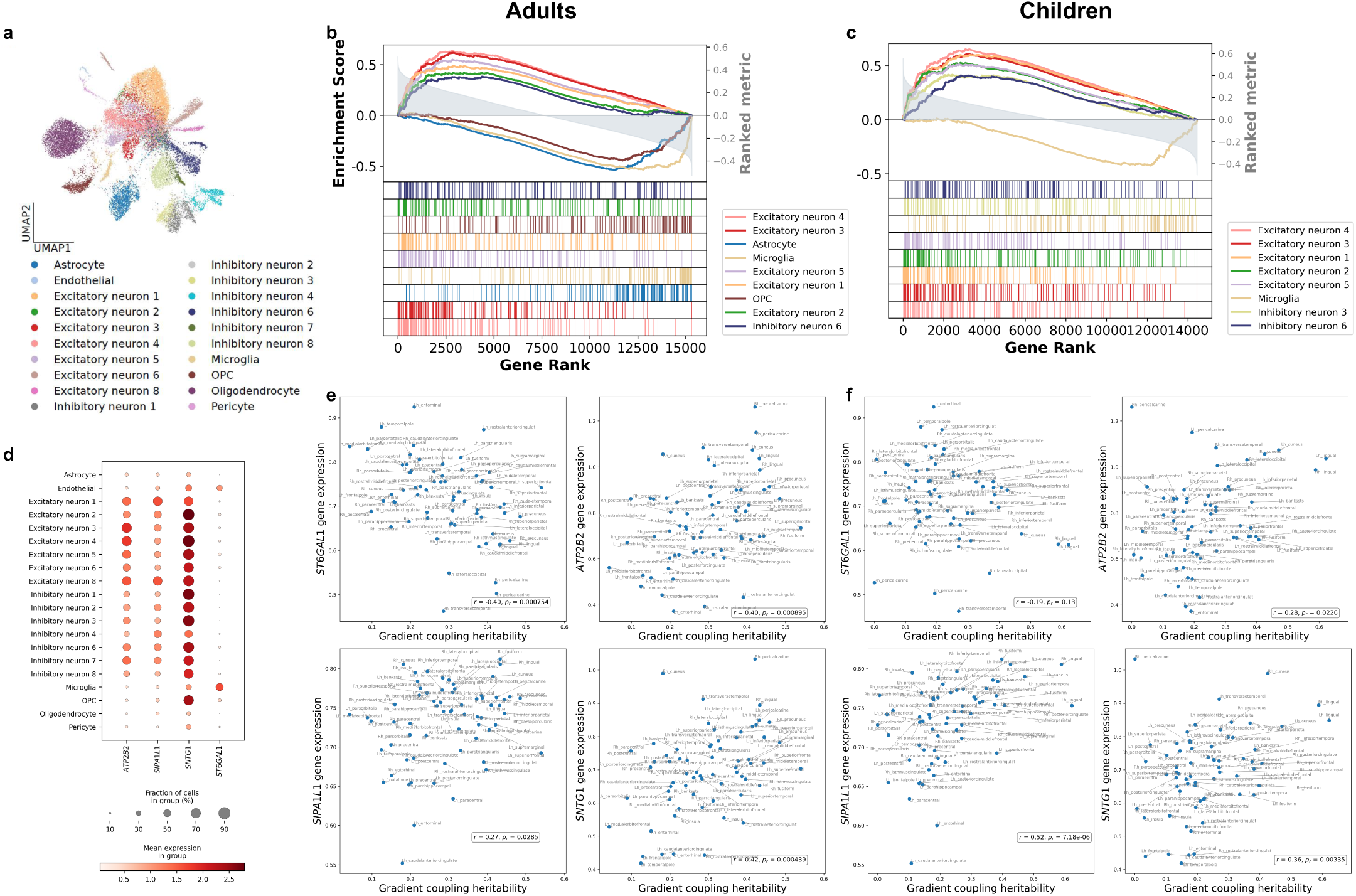
Imaging transcriptomics analyses of gradient coupling. **a** UMAP visualization [36] of the scRNA-seq data [37], with cells colored by annotated cell types. **b, c** GSEA results for the heritability of region-wise coupling between FCG1 and SCG1 in the two cohorts. Only cell types with absolute normalized enrichment scores greater than 2 are shown. **d** Dot plot of representative genes *ATP2B2*, *SIPA1L1*, *SNTG1*, and *ST6GAL1*. **e, f** Scatter plots showing associations between the heritability of D-K atlas-level gradient coupling (FCG1:SCG1) and expression levels of the genes in **d**, along with corresponding Spearman’s correlation coefficients (*r*) and p-values (*p_r_*).

To identify specific molecular contributors, we examined a set of genes highly expressed in the enriched excitatory neuron types and exhibiting strong positive correlations with gradient coupling heritability. Genes such as *ATP2B2*, *SIPA1L1*, and *SNTG1*, among the top neuron-enriched genes linked to heritable variation in gradient coupling, were both highly expressed and preferentially localized to excitatory neurons (Fig. 5d). In adults, *ATP2B2* showed Spearman’s *r* = 0.40*, p_r_* = 0.0090; *SIPA1L1* had *r* = 0.27*, p_r_* = 0.0285; and *SNTG1* showed *r* = 0.42*, p_r_* = 0.0004 (Fig. 5e). These genes were involved in calcium signaling and synaptic scaffolding [40, 41, 42], reinforcing the hypothesis that excitatory synaptic activity contributes to the spatial organization of heritable SF gradient coupling. In children, these genes also showed positive associations with gradient coupling heritability (Fig. 5f), albeit with weaker correlations for *ATP2B2* (*r* = 0.28*, p_r_* = 0.0226) and *SIPA1L1* (*r* = 0.52*, p_r_* = 7.18 × 10^−6^) and stronger correlation for *SNTG1* (*r* = 0.36*, p_r_* = 0.0034). While the magnitude of correlation varied across cohorts, the directionality of correlation was conserved with positive associations observed in both cohorts. This pattern indicated shared cellular drivers of genetic influence on gradient coupling that persist across development. Conversely, genes enriched in non-neuronal cell types, such as microglia, were negatively associated with gradient coupling heritability in both cohorts. For instance, *ST6GAL1*, a gene implicated in glycosylation and highly expressed in microglias, exhibited a strong negative correlation in adults (*r* = −0.40*, p_r_* = 0.0008), and a weaker yet still negative trend in children (*r* = −0.19*, p_r_* = 0.1300). These suggested potential opposing roles of glial and neuronal cells in shaping genetically influenced patterns of structural-functional alignment.

## Discussion

This work provided the first comprehensive study of brain SF gradient coupling and how it evolves over neurodevelopment and contributes to individual differences in brain organization and behavior. Prior research has primarily focused on single-modal gradients, with little attention to the cross-modal alignment between structural and functional gradients, and no systematic examination of its alterations over developmental stages, behavioral relevance, or genetic architecture. Here, we introduced SF gradient coupling as a neurobiologically grounded phenotype that quantifies the spatial alignment under various scales between structural and functional hierarchies in the human brain. Using multimodal neuroimaging data from two large-scale independent cohorts spanning childhood to adulthood, we demonstrated that SF gradient coupling captured key dimensions of neurodevelopmental reorganization and links macroscale brain architecture to cognition, behavior, and molecular mechanisms.

One of our central findings was the developmental transition in SF gradient coupling from childhood to adulthood. In children, functional organization was tightly coupled with principal structural axes, reflecting the early scaffolding role of white matter in shaping functional architecture. As maturation proceeded, this tight coupling gave way to a more differentiated and flexible structure and function relationship, characterized by a redistribution of alignment from primary to higher-order structural axes and increasing regional heterogeneity.

These dynamic changes in alignment underscored the need for a compact, interpretable framework capable of capturing both global and system-specific patterns of SF coordination. Crucially, our study addressed this need by introducing gradient coupling as a unified, cross-modal metric that quantifies the developmental refinement of brain cortical organization. This measure captures SF alignment across imaging modalities and spatial scales, providing a new framework for studying brain maturation.

Beyond developmental sensitivity, we also identified SF gradient coupling as a robust correlate of individual differences in behavior. The correspondence between gradient coupling and cognitive abilities was strongest during late childhood, particularly in females, and gradually attenuated in adulthood, mirroring the nonlinear trajectories of executive function and neural plasticity across development. In contrast, associations with sex and mental health became more prominent in adults. These patterns aligned with developmental models that posit adolescence as a critical window for emerging brain vulnerability architectures, during which deviations from normative coupling patterns may contribute to risk for psychopathology. In this context, gradient coupling bridges two key dimensions of neurodevelopment: maturational trajectory and interindividual variability, and may serve as a compact phenotype linking cortical wiring to diverse behavioral outcomes.

Mechanistically, the biological grounding of SF gradient coupling was supported by its heritability and cell-type specificity. Through classical ACE models, we found that heritability estimates were strongest among lower-order gradients and in cortical hubs such as the precuneus and superior frontal gyrus, highlighting a hierarchical genetic architecture of SF alignment. Spatially, heritability was highest in unimodal and heteromodal hubs with consistent bilateral organization across hemispheres. These brain regions aligned with known cortical hierarchies implicated in both global integration and cognitive control. At the cellular level, transcriptomic analyses revealed that the heritability of gradient coupling was enriched for excitatory neuronal signatures, particularly in deep-layer pyramidal neurons. This finding complemented recent evidence linking SF alignment to synaptic density and supported the critical role of excitatory circuits in shaping multimodal cortical gradients. In contrast, microglial signatures were negatively associated with coupling heritability, suggesting potentially antagonistic roles of glial and neuronal populations in mediating cross-modal gradient alignment.

Several implications follow from this work. First, SF gradient coupling may provide a new way to study developmental disorders that involve changes in brain structure or function, such as autism spectrum disorder, attention-deficit/hyperactivity disorder, and early-onset mood disorders [43]. Because it tracks normative age- and sex-related patterns of brain organization, it offers a framework for detecting atypical developmental trajectories that may signal increased risk for these conditions. Second, by demonstrating that coupling patterns shift from unimodal to transmodal systems with age, it highlights the importance of considering both regional and network-level differences in brain development. This could have practical value for precision psychiatry, where there is a need for brain-based markers that reflect age and network-specific changes [44].

While this study offers several novel insights, it also points to several directions for future research. First, our analysis was cross-sectional and age-stratified, limiting our ability to track changes within individuals over time. To better characterize how SF gradient coupling develops, future studies could resort to longitudinal designs that capture individual trajectories of brain gradient coupling changes. Second, although we employed standardized data processing pipelines across cohorts to improve comparability, differences in scanner protocols, sampling strategies, and behavioral assessments may still contribute to the observed differences between datasets. Future work using harmonized prospective datasets or advanced statistical harmonization techniques could further reduce these sources of variability. Lastly, while our use of cosine similarity between low-dimensional gradients under different scales provides an interpretable metric of SF gradient alignment, it may not capture the potential nonlinear relationship between gradients. Extending this framework to establish more rigorous manifold-based representations for coupling metrics, as well as to incorporate temporal fluctuations within fMRI time series and task-based modulations could yield a more comprehensive understanding of the dynamics and state-specific properties of SF gradient coupling.

## Methods

### Children cohort: ABCD study

The ABCD Study, funded by the National Institutes of Health, is a large-scale longitudinal research initiative investigating brain development and child health in approximately 11,880 children, recruited at ages 9–10 from 21 sites across the United States [31]. Employing comprehensive assessments, including advanced neuroimaging, genetic analysis, cognitive testing, mental health evaluations, and detailed environmental measures, the study tracks participants from childhood through adolescence into young adulthood. Its primary goals are to understand developmental trajectories, examine how genetic, behavioral, and environmental factors influence cognitive, emotional, social, and physical growth, and identify early indicators of mental health conditions to inform effective intervention strategies.

The neuroimaging and behavioral data used in this study were obtained from the NIMH Data Archive (NDA, https://nda.nih.gov/abcd). After rigorous quality control and preprocessing of the neuroimaging data, and exclusion of participants with incomplete covariate information, the final analytic sample consisted of 7,025 subjects (mean age = 9.9 years; 48.8% male). For the heritability analyses, we restricted the sample to 830 monozygotic (MZ) and dizygotic (DZ) twin pairs from 415 families, comprising 172 MZ pairs and 243 DZ pairs.

### Adults cohort: HCP study

The HCP is a major initiative in neuroscience, aimed at comprehensively mapping the neural pathways that underlie human brain function and elucidating the brain’s complex wiring architecture. A key component of this effort is the HCP Young Adult cohort (https://www.humanconnectome.org/study/hcp-young-adult/data-releases), which focuses on individuals aged 22 to 35—a life stage marked by relative brain maturity and stability, making it ideal for investigating normative brain structure and function. The HCP has established rigorous standards for data acquisition and preprocessing, providing an unprecedented view of the brain’s structural and functional connectivity. In addition to neuroimaging, participants complete a battery of behavioral and cognitive assessments, resulting in a rich dataset that relates brain connectivity to cognitive function and behavior. Neuroimaging data and most behavioral measures are publicly available at https://db.humanconnectome.org, subject to the HCP Open Access Data Use Terms.

Following quality control and data aggregation, our final analytic sample comprised 913 participants (mean age = 28.7 years; 46.55% male) from 414 families, including 111 MZ twin pairs, 59 DZ twin pairs, 511 full sibling pairs, and 95 singletons.

### Behavioral outcomes

For both children and young adults, we included the NIH Toolbox Cognition Battery, a standardized and developmentally appropriate assessment recommended for individuals aged 7 and older [45]. This battery spanned multiple cognitive domains, including executive function, episodic memory, language, working memory, processing speed, and sustained attention. It also included composite indices such as the Total, Fluid, and Crystallized Cognition scores, which provided aggregate measures of global, reasoning-based, and knowledge-based abilities, respectively. In our analyses, we included all seven individual task scores and the three composite scores to comprehensively assess the association between cognitive performance and the proposed coupling metric.

For mental health, we used instruments from the Achenbach System of Empirically Based Assessment: the Child Behavior Checklist (CBCL) for children and adolescents (ages 6–18), and the Adult Self-Report (ASR) for adults (ages 18–59). These parallel tools offered dimensional measures of behavioral and emotional functioning based on caregiver report (CBCL) and self-report (ASR), respectively. Specifically, we examined the broadband internalizing and externalizing scores, as well as key syndrome subscales including anxious/depressed, withdrawn/depressed, rule-breaking behavior, and aggressive behavior. A complete list of all cognitive and mental health measures used in our analyses, along with their corresponding variable codes, is provided in Supplementary Table 1.

### Multi-modal imaging acquisition, pre-processing and connectivity construction

The ABCD scan session followed a fixed sequence, beginning with a brief localizer for head alignment, followed by the acquisition of high-resolution 3D T1-weighted structural images, two runs of eyes-open resting-state fMRI, diffusion-weighted imaging, and 3D T2-weighted structural scans [31]. Depending on real-time motion monitoring, one or two additional resting-state fMRI runs were acquired to optimize data quality. The session concluded with task-based fMRI paradigms, including the Monetary Incentive Delay, Stop-Signal, and emotional n-back tasks, which probed reward processing, inhibitory control, and working memory. All data were collected using harmonized protocols across Siemens Prisma, GE 750, and Philips platforms at 21 U.S. sites, and underwent centralized quality control and Brain Imaging Data Structure-compliant preprocessing to ensure consistency and comparability across the cohort. Further details on the acquisition protocol can be found in [31].

In contrast, the HCP protocol was distributed across four approximately 1-hour sessions over two days, each beginning with a rapid localizer to ensure precise head positioning. Day 1 combined ultra-high-resolution structural imaging (T1- and T2-weighted) with alternating blocks of resting-state and task-evoked fMRI. Day 2 emphasized multi-shell diffusion acquisitions and included additional resting-state and task fMRI runs to enhance test–retest reliability [46]. All scans were performed on a customized Siemens Connectome Skyra with enhanced gradients and multiband EPI sequences. Data were processed through HCP’s Minimal Preprocessing Pipelines, including gradient nonlinearity correction, motion realignment, and Independent Component Analysis-based denoising, resulting in high-fidelity surface-based time series and diffusion models suitable for comprehensive connectomic analyses [47].

For both cohorts, we utilized preprocessed T1-weighted and diffusion MRI images, along with unprocessed resting-state fMRI data. Diffusion data were corrected for susceptibility-induced distortions, motion, and eddy currents, resampled to 1 mm isotropic resolution, and co-registered to the anatomical T1-weighted images to prepare for surface-based tractography [48]. T1-weighted scans were processed with FreeSurfer’s recon-all to reconstruct gray–white matter boundaries. Resting-state fMRI volumes underwent motion correction, bias-field correction, brain masking, and surface-based projection using FreeSurfer’s functional stream, followed by surface smoothing and nuisance regression (including global signal, motion parameters, and principal components from white matter and cerebrospinal fluid) [48].

Following these preprocessing steps, we constructed individual-level FC and SC matrices using the Surface-Based Connectivity Integration pipeline, a surface-based, atlas-free approach [48]. This approach used the white surface (the interface between white and gray matter) to construct connectivity matrices without being constrained by predefined brain parcellations [18].

According to [48], FC was obtained from resting-state fMRI by mapping blood oxygen level-dependent time series continuously onto each vertex of the reconstructed white–gray matter boundary in an atlas-free framework. Pairwise Pearson correlations between all vertex-wise time courses were computed and Fisher-z transformed to yield a dense, vertex-wise FC map that captured fine-grained synchrony across the cortical surface without reliance on prespecified parcellations. By treating FC as a continuous field on the white surface, this approach preserved the full topographic detail of functional network organization and facilitated direct, high-resolution comparisons with SC.

Building upon this continuous FC representation, SC was derived from diffusion MRI by estimating fiber orientation distributions via multi-tissue constrained spherical deconvolution and performing surface-enhanced tractography with seeds placed directly on the white-matter surface mesh. Each streamline’s endpoints were projected onto cortical vertices to construct a continuous density function of white matter connections, which was then smoothed and normalized to form a probabilistic, vertex-wise SC matrix perfectly aligned with the FC topology [48]. This continuous SC map provided a high-resolution anatomical scaffold that could be integrated seamlessly with the FC field for structure–function coupling analyses.

### Functional and structural gradient construction

Cortical gradients were extracted to capture macroscale organizational principles of structural and functional connectivity using the BrainSpace toolbox [49]. Specifically, we first thresholded the SC and FC matrices to retain the top 10% of connections, then transformed them into cosine similarity affinity matrices. Gradients were derived via diffusion mapping, a nonlinear dimensionality reduction technique known for its robustness to noise. In short, diffusion map embedding constructs a Markov operator *P* ^(^*^α^*^)^ = (Γ^(^*^α^*^)^)^−1^ *W* ^(^*^α^*^)^, where Γ^(^*^α^*^)^ is a diagonal matrix with 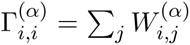, *W* ^(^*^α^*^)^ = Γ^−1^*^/α^ A* Γ^−1^*^/α^* is the affinity matrix *A* reweighted by the anisotropic diffusion parameter *α*, and Γ is the degree matrix of *A*. One then solves *P_α_ ϕ_ℓ_* = *λ_ℓ_ ϕ_ℓ_*, and defines the *m*-dimensional embedding at diffusion time *t* (the number of virtual steps in the random walk) as

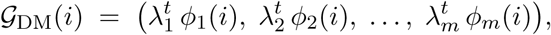

omitting the trivial *λ*_0_ = 1 eigenvector. Raising each eigenvalue to the *t*th power downweights modes that decay quickly. This yields gradients that capture both local and global connectivity structure with built-in noise robustness. In our implementation, we set *α* = 0.5 to balance the influence of sampling density, ensuring that neither dense nor sparse regions dominate the embedding. We used *t* = 0 to preserve the full spectrum of connectivity detail.

To enable group-wise comparisons, we aligned individual gradients by constructing a cohort-level template and applying Procrustes rotation [50], thereby minimizing inter-subject geometric variability. Following this pipeline, we retained the first 200 gradients per modality, which together explained over 99% of the variance in connectivity structure. For subject *i*, the resulting functional gradients 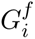 and structural gradients 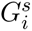 were matrices of dimension *V* × *D*, where *V* denotes the number of cortical vertices and *D* = 20 is the number of retained diffusion components, selected according to the highest eigenvalues in descending order.

In the cohort-level maps (Fig. 2a, g), the first two functional gradients accounted for 39% of variance in adults and 43% in children. In contrast, the first two structural gradients explained 49% of variance in adults but only 22% in children. While most information was captured by the first few components, variance explained declined gradually across higher-order gradients—dropping by less than 10% per gradient, and less than 5% beyond the fifth. To balance interpretability with information retention, we retained gradients that collectively explained at least 70% of the total variance. Specifically, in young adults, the top 20 gradients accounted on average for 74% of structural and 92% of functional connectivity variance. In children, the corresponding figures were 83% for structural and 70% for functional connectivity.

### SF gradient coupling

To quantify the alignment between SC and FC, we introduced SF gradient coupling, based on the gradients derived from each modality. We evaluated this coupling at two levels: (i) macroscale, capturing whole-brain gradient alignment, and (ii) subnetwork, assessing regional specificity (see Fig. 1 for illustration).

At the macroscale level, for each subject *i*, we computed the SF gradient coupling Ψ*_idd_*_′_ as the cosine similarity between the *d*th column of the functional gradient matrix 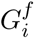 and the *d*^′^th column of the structural gradient matrix 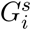, defined as:

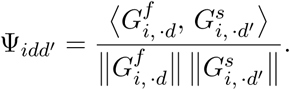

where 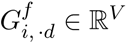 is the *d*th functional gradient for all *V* vertices; 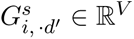 is the *d*^′^th structural gradient for all *V* vertices; *V* is the number of vertices in the whole brain; ⟨·, ·⟩ denotes inner product of two vectors; and ∥ · ∥ denotes the standard Euclidean norm of vectors. The resulting covariate vector **Ψ***_i_* = (Ψ*_i_*_11_, . . ., Ψ*_iDD_*) is of length *D*^2^, representing individual-specific SF gradient alignment. For example, selecting the top *D* = 20 gradients from each modality yields a coupling vector of length 400. In the following, we denote the dimension of SF gradient coupling vector as *d_sf_*.

To capture regional variation, we also constructed subnetwork-level SF gradient coupling, parcellating coupling signals according to two atlases: the Yeo 7-network functional parcellation [20] and the Desikan–Killiany (D-K) structural atlas [35]. Specifically, the gradients 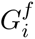 and 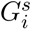 were first partitioned into *J* sub-vectors corresponding to the *J* predefined regions of interest (ROIs), denoted as 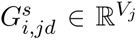 and 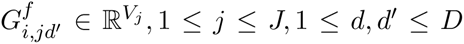, respectively. Here, *V_j_* is the number of vertices within the *j*th ROI. Then the subnetwork-level SF gradient coupling Ψ*_ijdd_*_′_ was calculated as the cosine similarity between each pair of functional and structural gradients within each of the *J* ROIs, defined as

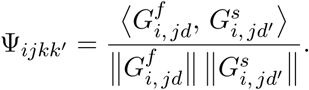

This yielded a subnetwork-level SF gradient coupling of dimension *d_sf_* = *J* × *D*^2^. For example, using the Yeo 7-network with *J* = 14 and *D* = 20 results in a *d_sf_* = 5, 600-dimensional vector per subject. These subnetwork couplings captured localized structure–function alignment within canonical brain systems.

### Associations between gradient coupling and behavioral outcomes

To examine associations between SF gradient coupling and behavioral outcomes, KRR and MLP models were applied independently to each SF gradient coupling–outcome pair, using either the macroscale or subnetwork-level gradient coupling vectors as input features.

In the KRR model [22], for each subject *i*, we denote the gradient coupling vector as **x***_i_*, and the outcome variable as *y_i_*. The model minimizes the regularized squared loss: 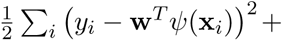 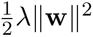, where **w** are the coefficients to be estimated; *ψ*(·) is a kernel function acting as a basis function; *λ* is a regularization parameter selected via cross-validation from the set {0.01, 0.1, 0.5, 1, 5, 10, 50, 10^2^, 10^3^, 10^4^, 10^5^}. Note that a higher *λ* would inflate the penalty term 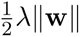 and lead to a more sparse model. Throughout our analysis, a linear kernel *ψ* was used.

For MLP, we first introduce the concept of neural networks. A neural network is a computational model inspired by the human brain’s structure, designed to recognize patterns, learn from data, and make predictions. It consists of layers of interconnected nodes or “neurons,” including an input layer that receives data, hidden layers that process it, and an output layer that delivers the final results. Each connection has associated weights and biases, which the network adjusts during training to optimize performance. Using activation functions, neural networks introduce non-linearity, enabling them to model complex relationships. The learning process involves forward propagation, where data flows through the network to produce an output, and backpropagation, where errors are used to update weights via optimization algorithms like gradient descent. In our analysis, we implemented MLPs with three fully connected layers for the prediction tasks. The input layer consists of the macroscale or subnetwork-level gradient coupling. The hidden dimension was set to 128, with a dropout rate of *p* = 0.5 to mitigate overfitting, and leaky ReLU activation with a negative slope of 0.2 was applied to the first two layers. For sex prediction, an additional Sigmoid function was included to map the outputs to the range [0, 1]. The number of training epochs was selected via cross-validation, with a maximum of 50.

### Model performance and feature importance

Model performance was evaluated using Pearson’s correlation coefficient for continuous variables and AUC for binary outcomes (e.g., sex). For robustness, we used 100 splitting replicates. The resulting 100 correlation coefficients or AUCs for each coupling-outcome correspondence were averaged or used for two-sample t-tests in the Results section. This procedure reinforced the robustness of the discovered association between the SF gradient coupling and outcomes of interest.

To assess feature relevance, we computed attribution scores for each model. For KRR, raw feature importance was computed as: 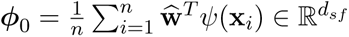, where *d_sf_* is the dimension of SF gradient coupling features, *d_sf_* = 400 when macroscale gradient couplings are used, *d_sf_* =5,600 when subnetwork-level gradient couplings under Yeo-7 network are used. For MLP, we used a saliency map [51], defined as the average gradient of the output with respect to the input features: 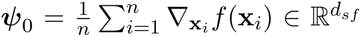, where ∇**_x_** *f* (**x***_i_*) indicates the partial derivative vector of the outcome of interest *f* (**x***_i_*) with respect to SF gradient coupling **x***_i_*. Raw importance values were then min–max normalized:

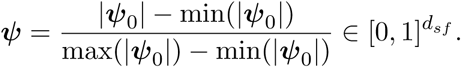

Higher normalized values indicated stronger predictive contributions of the corresponding features.

### Heritability estimation using the ACE model

We estimated the heritability of functional and structural gradients, as well as gradient coupling, using the classical ACE model based on twin and sibling data from the HCP and ABCD cohorts. The phenotype for each subject was modeled as:

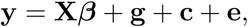

where **y** ∈ ℝ*^N^* is the phenotype vector for *N* individuals, **X** ∈ ℝ*^N^*^×^*^Q^* denotes the design matrix of fixed covariates (including age and sex), and ***β*** ∈ ℝ*^Q^* represents the corresponding fixed effects. The random effects include the additive genetic effect 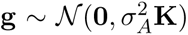, the shared environmental effect 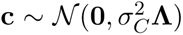, and the unique environmental effect 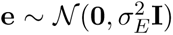. Here **K** is the genetic relatedness matrix and **Λ** represents the shared environment matrix, both constructed based on zygosity and parental identifiers. Heritability was defined as the proportion of phenotypic variance attributable to additive genetic effects:

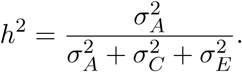

We used maximum likelihood estimation to fit the ACE model, where the variance components 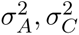, and 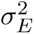 were estimated from the residuals of the fixed-effect model after regressing out the effects of age and sex. The ACE model was applied vertex-wise for functional and structural gradients, and region-wise for gradient coupling at both subnetwork (Yeo-7) and regional (D-K atlas) levels. For the young adult cohort, we used the full sample, which includes MZ and DZ twins, full non-twin siblings, and extended family structures sufficient for estimating genetic and environmental variance components. In contrast, for the children cohort, we restricted the heritability analyses to a genetically informative subset consisting only of same-sex MZ and DZ twin pairs (*n* = 830), rather than using the full sample of over 7,000 participants. This restriction was motivated by the extremely sparse genetic and shared environment matrices derived from the full sample, which would hinder the identifiability and stable estimation of variance components. By focusing on twin pairs with known zygosity, we ensured that the model assumptions were satisfied and that heritability could be robustly estimated.

### Imaging transcriptomics of SF gradient coupling heritability

To further investigate the biological mechanisms underlying the observed heritability patterns in functional gradients, structural gradients, and their coupling, we performed an imaging transcriptomic analysis that integrated regional heritability estimates with gene expression profiles from AHBA [38] and cortical single-cell transcriptomics data [37]. Following the established imaging transcriptomics workflow [52], we extracted region-wise gene expression profiles from the AHBA using the *abagen* toolbox [53], mapped to the D-K atlas [35]. Gene expression values were normalized within each region to a total count of 10,000. We computed Spearman’s rank correlations between the heritability values and the expression of 15,633 genes across 66 cortical ROIs (two ROIs with missing gene data were excluded), and ranked genes in descending order based on their correlation with the phenotype-specific heritability pattern. To assess cell-type specificity, we performed GSEA using the *GSEApy* library [39]. Cell-type–specific gene sets were derived from scRNA-seq data [37], where the top 200 differentially expressed genes for each cell type were identified using *t*-tests implemented via the *SCANPY* pipeline [54]. GSEA was applied to the ranked gene list for each phenotype (e.g., heritability of the first functional gradient or gradient coupling), and only cell types with an absolute normalized enrichment score (NES) greater than 2.0 were reported.

### Analysis robustness and stability

Since we obtained 100 realizations of gradient coupling-outcome associations and 100 normalized feature importance scores for each gradient coupling–outcome pair, these replications enabled us to perform two-sample t-tests and calculate mean and standard deviation summaries in downstream analyses. This procedure ensured the robustness of the gradient coupling-outcome correspondence and mitigated potential issues of overfitting and sampling bias.

The two predictive modeling approaches, KRR and MLP, demonstrated strong concordance. In particular, the overall Pearson’s r between KRR and MLP in the gradient coupling-behavior association was 0.94, highlighting the robustness of the associations we identified. We presented the association results based on MLP in Supplementary Fig. 2. When compared to Fig. 3a using KRR, MLP produced highly similar association patterns, further validating the robustness of the analysis. Additionally, KRR and MLP exhibited similar patterns of feature importance. Examples illustrating this similarity were provided in Supplementary Figs. 3 and 4. The averaged feature importance values, computed across both methods, were reported in the Results section.

## Data Availability

All data produced in the present work are contained in the manuscript.

https://abcdstudy.org

https://www.humanconnectome.org

## Data availability

Neuroimaging and behavioral data from the ABCD Study can be obtained via the NIH Data Archive (NDA; https://nda.nih.gov/abcd) with approval from the ABCD consortium. Neuroimaging data and most behavioral measures from the HCP are publicly available at https://db.humanconnectome.org; access to restricted data is subject to approval.

## Code availability

The data preprocessing software FreeSurfer v6.0 is available at https://surfer.nmr.mgh. harvard.edu/. The Surface-Based Connectivity Integration pipeline can be accessed at https://github.com/sbci-brain/SBCI_Pipeline, and the BrainSpace toolbox is available at https://github.com/MICA-MNI/BrainSpace/tree/master. Python 3.8 was used for data processing and analysis. Code for gradient coupling construction, association analyses, and heritability estimation is publicly available at https://github.com/Naomi-Ding/SF-Gradient-Coupling.

## Acknowledgment

This research was partially supported by National Institutes of Health (NIH) grants R01AG068191, RF1AG081413 and R01EB034720. We thank the individuals represented in the ABCD and HCP studies for their participation and the research teams for their work in collecting, processing and disseminating these datasets for analysis.

Some data used in the preparation of this article were obtained from the ABCD study, held in the National Institute of Mental Health Data Archive. This is a multisite, longitudinal study designed to recruit more than 10,000 children age 9–10 years and follow them over 10 years into early adulthood. The ABCD study is supported by the NIH and additional federal partners under award numbers U01DA041022, U01DA041028, U01DA041048, U01DA041089, U01DA041106, U01DA041117, U01DA041120, U01DA041134, U01DA041148, U01DA041156, U01DA041174, U24DA041123, U24DA041147, U01DA041093 and U01DA041025. A full list of supporters is available at https://abcdstudy.org/federal-partners.html. A listing of participating sites and a complete listing of the study investigators can be found at https://abcdstudy.org/scientists/workgroups/. ABCD consortium investigators designed and implemented the study and/or provided data but did not necessarily participate in analysis or writing of this report. All procedures in the ABCD study were approved by the institutional review boards at ABCD collection sites (approval numbers 201708123 and 160091).

HCP data were provided by the HCP, WU-Minn Consortium (principal investigators D. Van Essen and K. Ugurbil; 1U54MH091657) funded by the 16 NIH institutes and centers that support the NIH Blueprint for Neuroscience Research and the McDonnell Center for Systems Neuroscience at Washington University. All experimental procedures in the HCP study were approved by the institutional review boards at Washington University (approval number 201204036).

## Author contributions

Y.Z., S.G., Z.G., and S.D. conceptualized and designed the study. Z.Z. collected and processed the neuroimaging data. S.G., Z.G., S.D., and G.W. analyzed the data. S.G., Z.G., and S.D. wrote the original draft and revised the manuscript. All authors edited and reviewed the final manuscript.

