## Supplementary Materials for "Brain Functional-Structural Gradient Coupling Reflects Development, Behavior and Genetic Influences"

### Supplementary Material for "Brain Functional-Structural Gradient Coupling Reflects Development, Behavior and Genetic Influences"

Simiao Gao[1,†], Zhiling Gu[1,†], Shengxian Ding[1,†], Gefei Wang[1], Zhengwu Zhang[2], Hongyu Zhao[1] & Yize Zhao[1,*]

[1] Department of Biostatistics, Yale University, New Haven, CT 06510, USA
[2] Department of Statistics and Operations Research, University of North Carolina at Chapel Hill, Chapel Hill, NC 27599, USA
[†] These authors contributed equally to this work

Supplementary Table 1: Cognitive and Mental Health Measures with Corresponding Codes in ABCD and HCP Studies.

| Domain | Outcomes | ABCD Codes | HCP Codes | Outcome Dictionary |
|---|---|---|---|---|
| Mental Health | Internal | cbcl_scr_syn_internal_t | ASR_Intn_T | Internalizing Problems |
| | External | cbcl_scr_syn_external_t | ASR_Extn_T | Externalizing Problems |
| | Anx/Dep | cbcl_scr_syn_anxdep_t | ASR_Anxd_Pct | Anxious/Depressed Problems |
| | With/Dep | cbcl_scr_syn_withdep_t | ASR_Witd_Pct | Withdrawn/Depressed Problems |
| | RuleBreak | cbcl_scr_syn_rulebreak_t | ASR_Rule_Pct | Rule-Breaking Behavior |
| | Aggressive | cbcl_scr_syn_aggressive_t | ASR_Aggr_Pct | Aggressive Behavior |
| Cognition | PicVocab | nihtbx_picvocab_agecorrected | PicVocab_AgeAdj | Picture Vocabulary |
| | Flanker | nihtbx_flanker_agecorrected | Flanker_AgeAdj | Flanker Inhibitory Control and Attention |
| | ListSort | nihtbx_list_agecorrected | ListSort_AgeAdj | List Sorting Working Memory |
| | CardSort | nihtbx_cardsort_agecorrected | CardSort_AgeAdj | Dimensional Change Card Sort |
| | ProcSpeed | nihtbx_pattern_agecorrected | ProcSpeed_AgeAdj | Pattern Comparison |
| | PicSeq | nihtbx_picture_agecorrected | PicSeq_AgeAdj | Picture Sequence Memory |
| | ReadEng | nihtbx_reading_agecorrected | ReadEng_AgeAdj | Oral Reading Recognition |
| | CogFluid | nihtbx_fluidcomp_agecorrected | CogFluidComp_AgeAdj | Fluid Composite |
| | CogCrystal | nihtbx_cryst_agecorrected | CogCrystalComp_AgeAdj | Crystallized Composite |
| | CogTotal | nihtbx_totalcomp_agecorrected | CogTotalComp_AgeAdj | Total Cognition Composite |

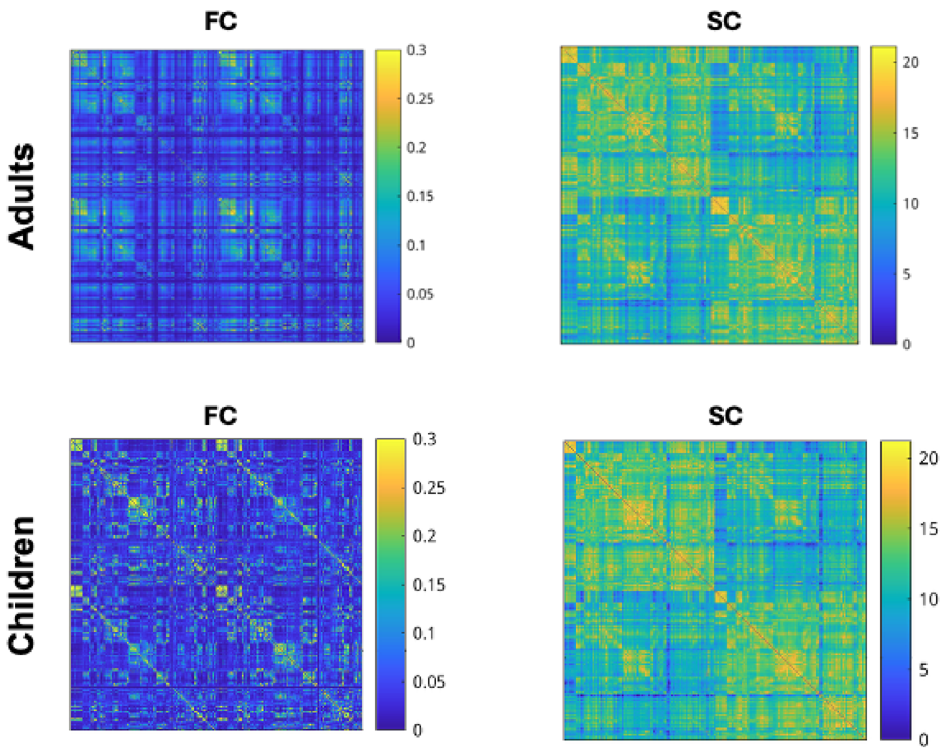


Supplementary Figure 1: Group-level functional and structural connectivity of young adults and children after Procrustes transformation. FC: functional connectivity; SC: structural connectivity.

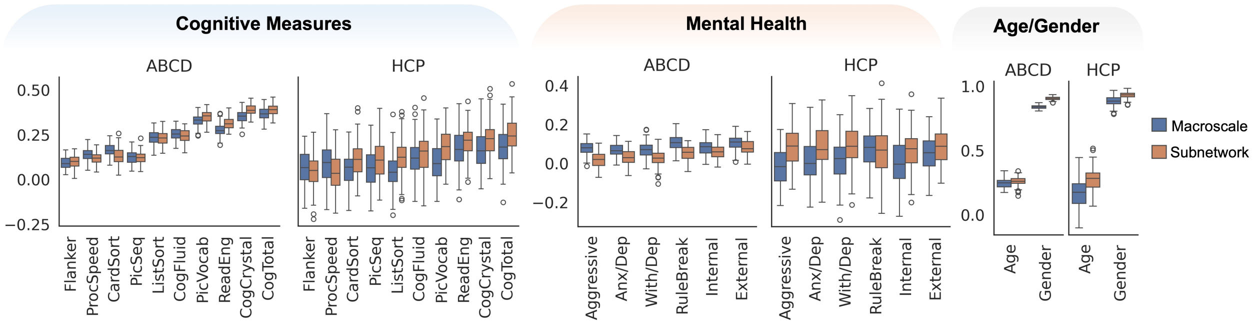


Supplementary Figure 2: Boxplots illustrating the association between gradient coupling and behavioral outcomes at the macroscale (blue) and Yeo-7 subnetwork level (orange) using MLP. Correlation coefficients are shown for continuous measures (cognitive and mental health), and AUC values for age and gender.

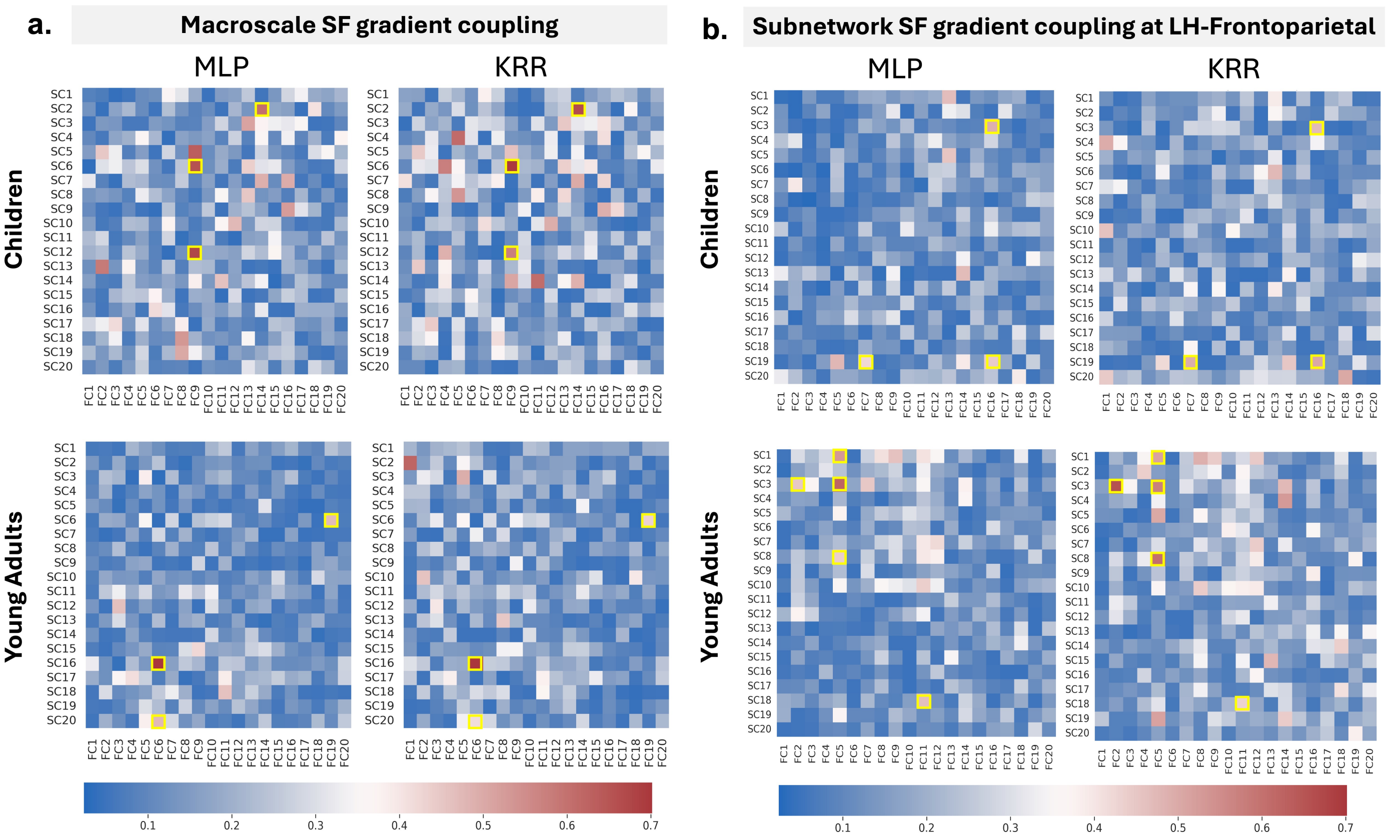


Supplementary Figure 3: Examples of feature importance patterns of SF gradient couplings in predicting cognitive outcomes. **a** feature importance of macroscale SF gradient couplings **b** feature importance of subnetwork SF gradient couplings in the left hemisphere (LH) frontal-parietal region.

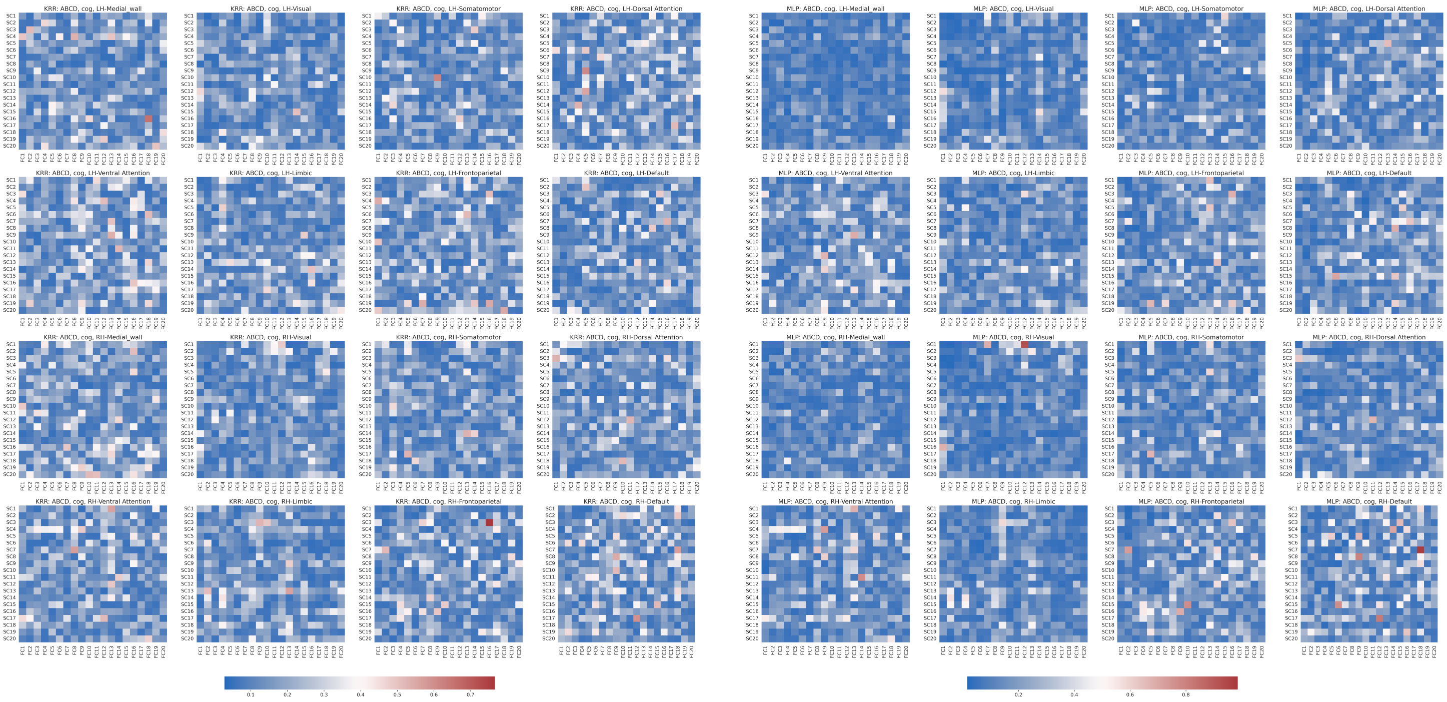


**(a)** Children, KRR (Left) vs MLP (Right)

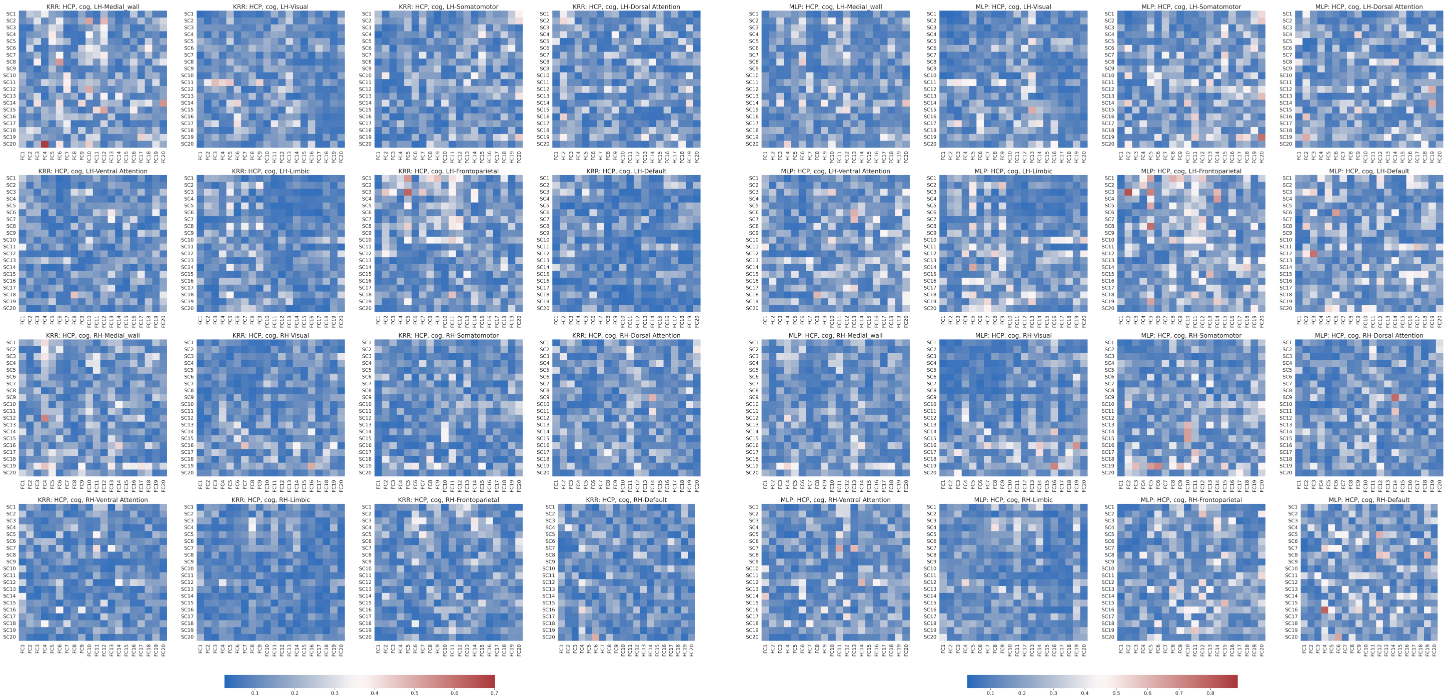


**(b)** Young Adults, KRR (Left) vs MLP (Right)

Supplementary Figure 4: Feature importance map for predicting cognitive outcomes using the subnetwork SG gradient coupling for Children (**a**) and Young Adults (**b**).

### Supplementary Results

#### Heritable Structure of Functional and Structural Gradients

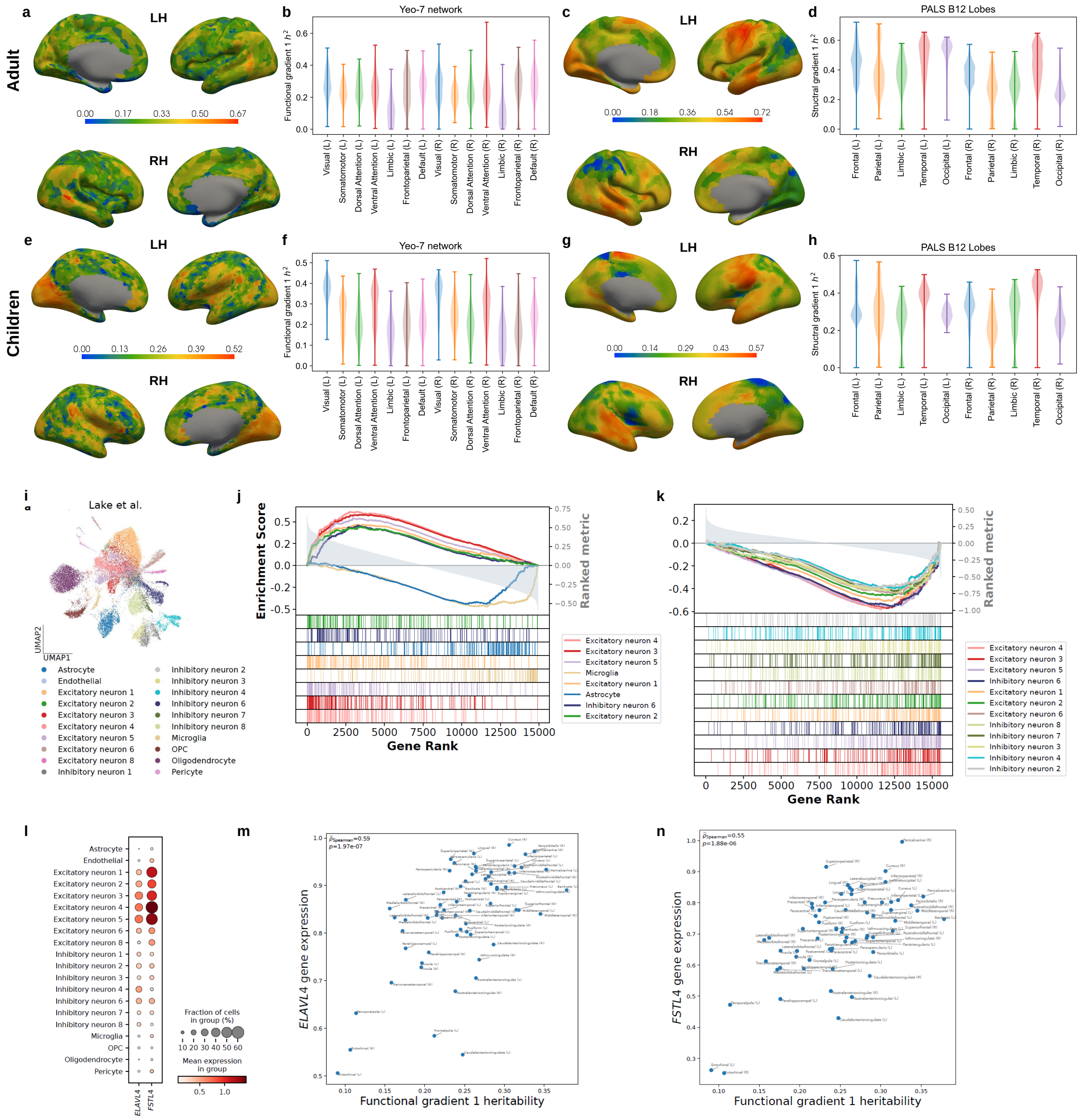

**Supplementary Figure 5: Heritability & imaging transcriptomics analyses of functional and structural gradients.** **a, e**. Heritability estimates of the first functional gradient for HCP & ABCD study. **b, f**. Heritability estimates of the first functional gradient grouped by Yeo-7 networks. **c, g**. Heritability estimates of the first structural gradient. **d, h**. Heritability estimates of the first structural gradient grouped by PALS B12 lobes. **i**. Two-dimensional visualization of the single-cell RNA-sequencing dataset reference [1] using UMAP [2]. Cells are colored by their cell type labels. **j, k**. GSEA results of the first functional & structural gradient heritability. Only cell types with absolute values of normalized enrichment scores greater than 2 are shown. **l**. Dot plot of genes *ELAVL4* and *FSTL4*. **m, n**. Scatter plots showing the relationship between the first functional gradient heritability and gene expression values of *ELAVL4* (**m**) and *FSTL4* (**n**).

To contextualize the coupling between functional and structural gradients, we first estimated the vertex-wise heritability of functional and structural gradients using the ACE model applied to monozygotic and dizygotic twin pairs in both the HCP and ABCD cohorts. Functional gradients exhibited moderate heritability, with mean $h^2$ estimates of $h^2 = 23.49\%$, $20.86\%$, and $17.83\%$ for the first three gradients in HCP, and similar values in ABCD (23.97%, 23.28%, and 17.51%, respectively). In contrast, structural gradients were consistently more heritable, particularly in HCP, where the first gradient reached a mean heritability of 48.12%, compared to 32.16% in ABCD, with notably lower heritability. To visualize the spatial distribution of heritability estimates for both modalities, we mapped the first functional and structural gradients onto the cortical surface (Supplementary Fig. 5 **a, c, e, g**), showing widespread and topographically organized heritability patterns across the cortical surface for both datasets. Structural gradients consistently exhibited stronger and more localized heritability, particularly in occipital regions and other posterior cortices, including posterior parietal and temporal areas. Inter-gradient heritability comparisons further revealed strong correlations among functional gradients (Pearson's $r = 0.44 - 0.51$, $p < 10^{-100}$), suggesting shared genetic influences. In contrast, the heritability of structural gradients was more distinct from one another, with weaker correlations (maximum $r = 0.15$, $p < 10^{-19}$), pointing to more heterogeneous genetic underpinnings. To explore functional specificity, we parcellated the first functional gradient heritability by Yeo-7 networks (Supplementary Fig. 5 **b, f**). This analysis revealed symmetrical hemispheric patterns and consistently reduced heritability within the limbic system, possibly reflecting a greater influence of environmental factors on regions subserving emotion and memory [3, 4]. By contrast, higher heritability was observed in transmodal systems, particularly the frontoparietal and default mode networks, especially in the right hemisphere. We then examined the heritability of the first structural gradient across cortical lobes using the PALS-B12 atlas (Supplementary Fig. 5 **d, h**). While structural heritability was evident across all lobes, greater hemispheric asymmetries were observed than for functional gradients. For instance, in both datasets, the left parietal lobe showed higher heritability than its right-hemisphere counterpart. In HCP, the right frontal and temporal lobes also displayed stronger heritability than the parietal lobe, whereas in ABCD, structural heritability tended to be higher in the left hemisphere overall.

To probe the cellular basis of these heritability patterns, we conducted an imaging transcriptomics analysis using data from the Allen Human Brain Atlas [5] and cortical single-cell RNA sequencing (scRNA-seq) datasets [1]. The scRNA-seq reference dataset includes 20 brain cell types, allowing us to examine spatial relationships between brain phenotypes and cell-type activities (Supplementary Fig. 5 **i**). We focused on the first functional gradient in HCP due to its high heritability and cross-gradient consistency. By correlating the heritability profile across 66 cortical ROIs with expression levels of 15,633 genes, we identified neuronal genes most strongly associated with heritable variation in the first functional gradient. Gene set enrichment analysis (GSEA, [6]) revealed significant enrichment of excitatory neuron subtypes, particularly clusters labeled as excitatory neurons 3–5 (Supplementary Fig. 5 **j**). Notably, genes such as ELAVL4 and FSTL4—highly expressed in cortical layers 4–6—showed strong positive correlations with functional gradient heritability (Spearman's $\rho = 0.59$, $p = 1.97 \times 10^{-7}$; and $\rho = 0.55$, $p = 1.88 \times 10^{-6}$; Supplementary Fig. 5 **l-n**). In contrast, non-neuronal cell types, including astrocytes and microglia, showed enrichment for genes with negative correlations, highlighting potential opposing influences on heritable functional organization (Supplementary Fig. 5 **l**). For the first structural gradient, transcriptomic associations were generally weaker and skewed away from neuron-enriched genes (Supplementary Fig. 5 **k**), suggesting distinct cellular drivers of genetic influence on structural versus functional gradients.

Together, these results confirm that functional and structural gradients are heritable traits with distinct genetic and cellular architectures. The functional gradients appear more tightly coupled to excitatory neuronal gene expression, whereas structural gradients reflect more diverse and spatially asymmetric genetic influences. These findings provide a strong rationale for investigating gradient coupling as a biologically meaningful and heritable marker of integrated brain organization across modalities.
